# Synergistic Torso-Muscle-Controlled Detached Robotic Hand: A Novel Approach for Post-Stroke Hand Rehabilitation

**DOI:** 10.1101/2025.05.02.25326083

**Authors:** Joshua N. Posen, Joshua Lee, Frank L. Hammond, Stephen N. Housley, Andrew J. Butler, Minoru Shinohara

## Abstract

This study aimed to develop a novel rehabilitative approach for post-stroke hand movement using a simple detached robotic hand and synergistic torso muscle activities for reaching and to perform a pilot test on its functionality and feasibility. In reference to a mental practice that does not activate hand muscles, enhanced cognitive engagement would be achieved without hand activation using the externally present, visible, and audible robotic hand by activating the non-hand muscles associated with hand function. A simple and low-cost robotic hand was developed and placed distal to the hidden resting hand as if it were a functional extended hand. The opening and closing motions of the detached robotic hand were controlled by electromyogram of the anterior and posterior torso muscles associated with reaching and retrieving while providing visual and auditory feedback. The functionality of the developed system was confirmed on the repeatability of the range of duration, excursion, and response time with low variability within an acceptable range. An able-bodied adult and five mildly impaired stroke survivors embodied the detached robotic hand by successfully controlling it with or without concurrent testing of their biological finger. In the concurrent finger tests, increased reactive force and hand muscle activity were observed in most participants. These observations confirmed that the developed approach that controls a detached robotic hand with reaching-associated torso muscles is functional and applicable to stroke survivors with and without involving the biological human hand. The robotic hand system detached from the user and controlled by the voluntary effort of their reaching-associated torso muscles has enabled future studies to examine the efficacy of synergistic muscle-robot interaction as a potential rehabilitation tool.

## Introduction

Upper limb motor function is often impaired due to neurologic injuries such as stroke and spinal cord injury. In the United States alone, more than 795,000 people suffer a stroke every year [4], and approximately 60% of stroke survivors experience substantial impairments in hand function [3], requiring long-term rehabilitative therapy [10]. Few interventions improve their upper-limb function. Conventional motor training involves repeatedly exercising the affected hand, but it is often difficult to activate the impaired hand in a functional manner. To facilitate the movement, various robotic rehabilitation systems have been developed, such as the Exo Glove Poly, iHandRehab, IntelliArm, HandSOME, OHAE, and Hand of Hope, in which a robot is attached to the limb or hand to provide physical assistance or resistance [2,6,16,18,19,27]. However, patients can only execute and observe their *incomplete* hand movement because the speed and range of hand motion with an attached robotic system are still limited due to the curled and stiffened posture of their affected hand (Fig. 1C). Observations of actions with limited range of motion and with attached robotic equipment are not optimal for cognitive or neural engagement because of the discrepancies from the intended and expected shape and movement during exercise. Hence, it is imperative to develop a new modality of robotic rehabilitation that can overcome these limitations. Our novel and unique idea is to develop a robotic hand rehabilitation system that utilizes a “detached” robotic hand, which resolves the discrepancies across the intention, execution, and observation of a “hand” movement.

**Fig. 1.**
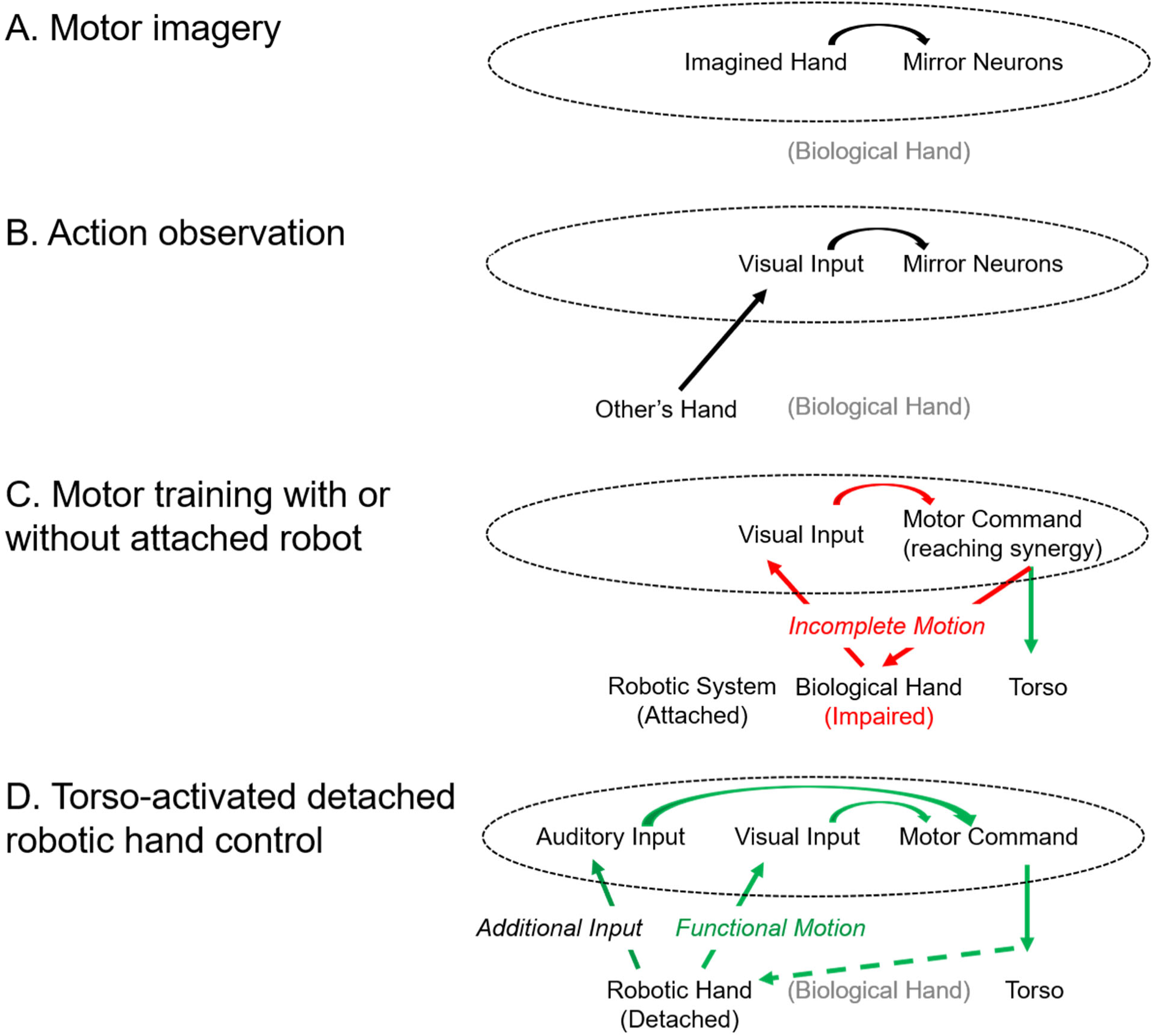
A schematic depicting the pathways activated by different rehabilitation approaches and the integration of external stimuli. Motor imagery (a) and action observation (b) are performed without physical motion of the biological hand but engagement in regions of the brain (*in dotted ovals*) for motor command is present. Robotic rehabilitation systems which attach to the impaired biological hand result in incomplete motion and its observation (*in red*), affected by muscle stiffness and posture of the impaired hand, which can influence motor command. A robotic system with a detached hand (d) enables the observation and creation of functionally correct motion (*in green*) through reaching synergy muscles (torso) with additional auditory input (*in green*) regardless of the functional capability of the biological hand, which can positively influence motor command.

Action observation and motor imagery have been leveraged as mental practice in stroke rehabilitation, especially for the upper limb, to enhance embodiment and recruit the mirror neuron system (Figs. 1A & 1B). The mirror neuron system is engaged when observing or imitating a physical motion. In mental practice for stroke rehabilitation, an affected limb is not physically involved, but the associated and additional neural areas in the brain can be active, including the mirror neuron system [5,8,35,37]. Repetitions of such neural activation and its combination with other types of rehabilitation (e.g., motor training) can facilitate adaptations in those neural areas [20,23], leading to motor recovery. The problem is that mental practice is difficult to engage and perform because it is executed internally without a corresponding movement of a visible hand. To resolve discussed limitations, we decided to develop a novel neurorehabilitation system that utilizes the observation and active control of the opening and closing of a detached robotic hand via synergistic (*i.e.*, associated with specific hand movements) proximal muscle activation that does not require the involvement of their biological hand (Figs. 1D & 2).

**Fig. 2.**
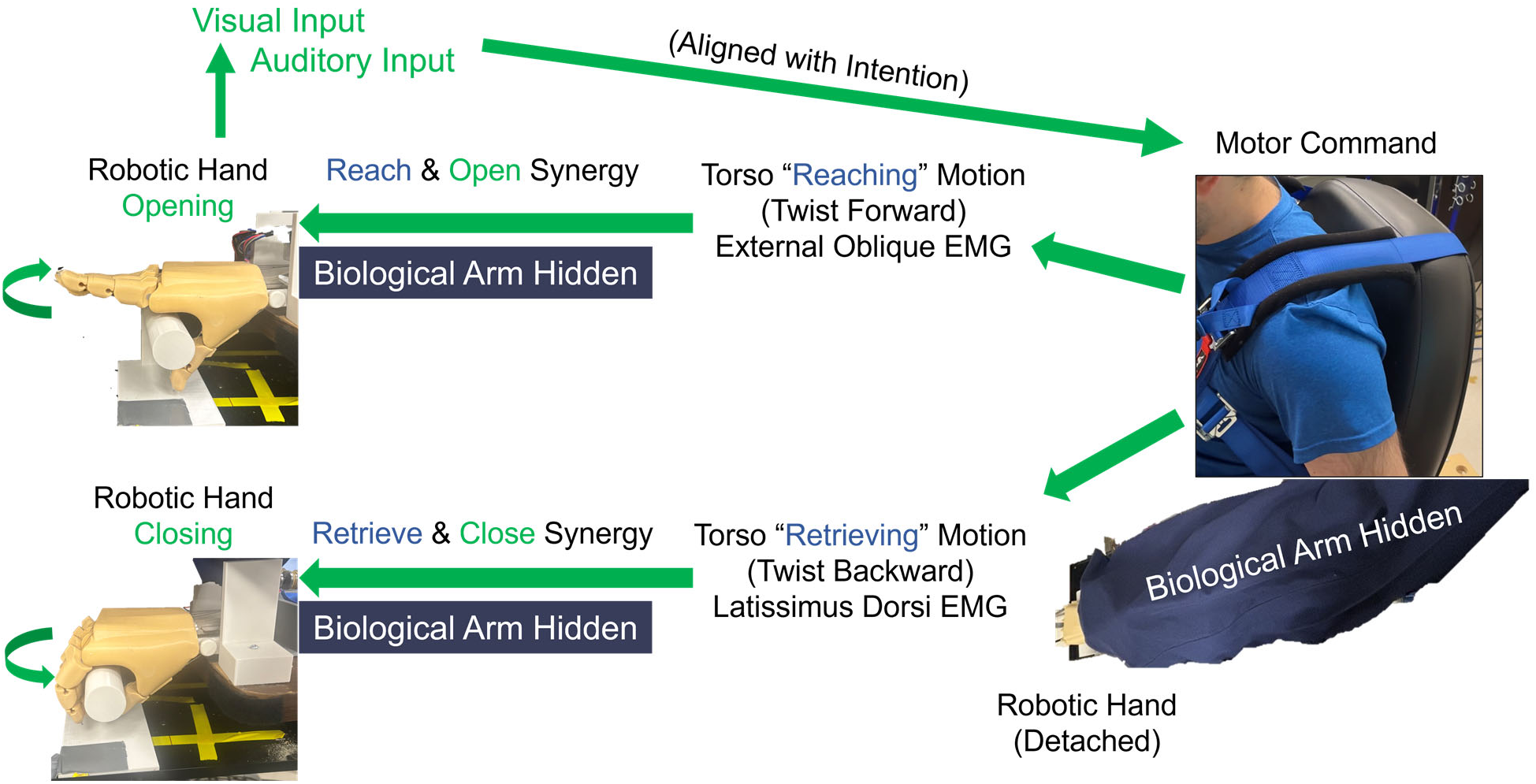
Overview of the novel application of the “detached” robotic hand as an extension of the hidden limb for rehabilitation. A user, restrained by a harness, produces motor command to activate synergistic torso muscles during mimicked reach and hand opening (external abdominal oblique, *top*) and retrieve and hand closing (latissimus dorsi, *bottom*) tasks, which controls the robotic hand. The user observes the robotic hand and receive visual and auditory input that is aligned with the intention.

In reaching and retrieving an object, the anterior (e.g., external abdominal oblique) and posterior (e.g., latissimus dorsi) torso muscles for leaning forward and backward are functional synergistic muscles that accompany their hand’s opening-reaching and closing-retrieving motions, respectively. Additionally, stroke survivors often rotate the torso inward and backward in reaching and retrieving, respectively [9,24,28]. As an alternative to trying to control the impaired biological hand, the mimicked synergistic interaction with a robotic hand placed distal to their hidden biological hand is expected to induce an illusion as if the robotic hand is a part of their body extended from their limb (Fig. 2). It can facilitate the involvement of neural pathways for the non-activated distal muscles due to cognitive engagement with the externally present, visible, and audible robotic hand. The rationale includes proven procedures that can facilitate neuromotor excitability: concurrent motor imagery and action observation [29,36], concurrent visual and auditory inputs [1], and activation of the synergistic proximal muscles associated with distal muscles [11,15,32]. The unique integration of the evidence-based paradigm would benefit neuromotor recovery for distal muscles. The development of this system enables studying scientific and clinical inquiries as to the efficacies and mechanisms of this integrated paradigm in stroke rehabilitation by utilizing a detached robotic hand.

The exploration into using a detached robotic hand in rehabilitation is possible at a low cost with the increased prevalence of 3D printing for research prototyping. To assess the feasibility of a detached robotic hand as a rehabilitation tool, we need first to create a proof-of-concept system for human studies. Therefore, we developed a simple, low-cost (< US$150), yet reliable system for human interaction in this study. The system is limited to 1 DOF, opening and closing of the hand, and operates through an on-off control scheme to generate the motion within a defined duration of time. Potentially, a non-immersive virtual reality, i.e., a virtual hand on a 2-dimensional video space on a monitor, could be a simple, low-cost alternative to the detached robotic hand. However, observing a 2-D virtual hand movement on a monitor can limit the depth perception and spatial awareness by the user, which can attenuate the sense of presence, involvement, and embodiment. Indeed, the degree of realism, including 3D vs. 2D, is known to influence the modulation of the mirror neuron system. For example, the kinematics observed when completing a reaching task in a virtual space is seen to be significantly different than the motion observed in the physical world [14,21,33]. fMRI studies have shown that observing 2D actions evokes weaker responses in the mirror neuron system compared with observing 3D actions [13,17]. Hence, in this study, we aimed to develop a detached robotic rehabilitation system and perform a pilot test on a young, able-bodied adult to demonstrate the proof of concept and five stroke survivors to evaluate the feasibility. The contribution of this study includes the unique idea of resolving the discrepancies in hand rehabilitation, the low-cost realization of the idea for conducting future scientific and clinical studies, and the demonstration of the feasibility. A preliminary version of this work has been reported in an abstract form [25, 26].

## Methods

Our unique idea is to have a human control and observe the intended motions of a detached anthropomorphic robotic hand via the torso activity synergistically associated with biological hand motions during hand opening-reaching and hand closing-retrieving motions, respectively (Fig. 2). The robotic hand was 3D-printed for a standard adult size with two functional movements: the opening (i.e., synergistic hand opening during reaching) and closing of the fingers (i.e., synergistic grasping and holding during retrieving) (Fig. 3). A user controls and observes these motions by activating their anterior (external abdominal oblique) and posterior (latissimus dorsi) torso muscles for mimicked reaching and retrieving motions, respectively.

**Fig. 3.**
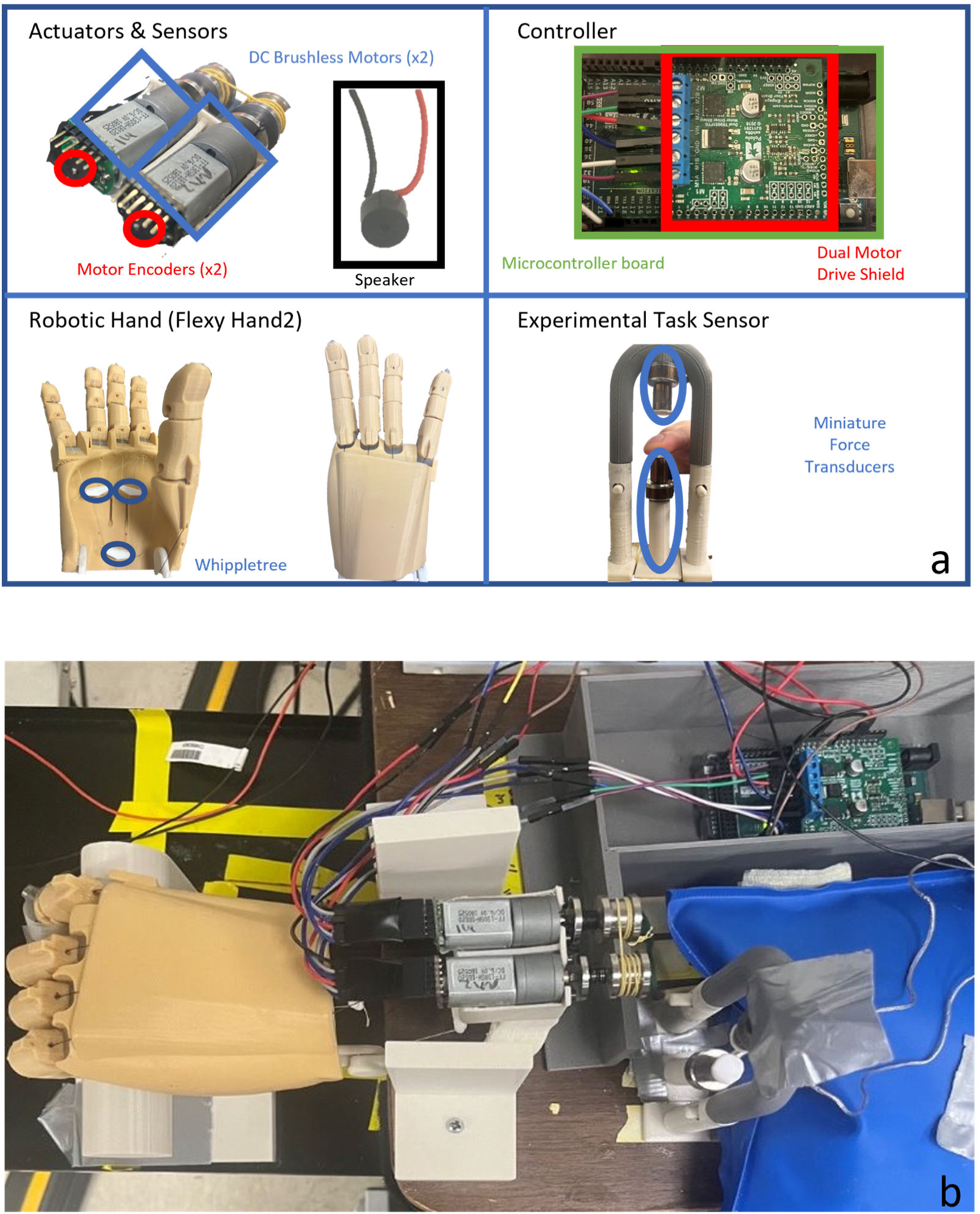
System breakdown (a) and photo (b) of the experimental setup for operation of the detached robotic hand. The detached robotic hand is actuated by two DC motors. The motors are powered and controlled by a microcontroller and specialized shield. The motor position is sensed by attached motor encoders. Force transducers are present to test the feasibility of action by the biological hand while interacting with the robotic hand. A speaker is positioned near the robotic hand to generate sounds to indicate the orientation of the robotic hand.

Our initial design considered the post-stroke population coupled with tasks without hand muscle activation. In human testing, all subjects were right-handed, and the additional inclusion criteria for the post-stroke subjects included 21-75 years old, longer than six months post-stroke, persistent hemiparesis on the right upper extremity (UE), residual UE voluntary movement as indicated by a score of 1-3 on the motor arm item of the NIH Stroke Scale (NIHSS) and a score of 19-55 on the UE portions of the Fugl Meyer Assessment, and preserved cognitive function and ability to follow and read simple instructions as indicated by a score of ≥ 1 on item #9 on the best language items of the NIHSS. There were several exclusion criteria for post-stroke subjects, including limited active range of finger motion < 10 degrees in flexion and extension, inability to activate trunk muscles, severe sensory impairment, excessive pain in any joint, and cognitive impairment. None of the post-stroke subjects participated in synchronous rehabilitation or research activities. The screening on these criteria was conducted by one of the co-authors with a physical therapy licensure in the State of Georgia (SNH). The experimental procedure was approved by the Institutional Review Board of the Georgia Institute of Technology (H21154). The study relied on subjects recruited as part of a clinical trial (NCT04962698).

### *A.* Intended Functional Characteristics

The fundamental characteristics we considered with the robotic hand include the duration, range of motion, and response time of the digit motions associated with reaching and retrieving in stroke survivors. Humans naturally open their hand while reaching for an object and close the hand during retrieving. The corresponding durations are, on average, ∼1 s and 1.13 s in able-bodied populations, 1.88 s and 1.68 s in mild impairment, and 2.59 s and 2.21 s in moderate impairment with stroke [7]. Hence, we targeted the 1.5-2 s range for the proof-of-concept study with stroke survivors to appropriately align with their anticipated movement patterns. The range of motion for digit flexion and extension is impaired following stroke, with the metacarpophalangeal joint of the index finger limited to ∼20° [22]. In this study, we aimed for the robotic hand to cover the range of motion of average able-bodied adult males to grasp a door handle, i.e., ∼45° [22]. The response time was between the torso muscle electromyogram (EMG) crossing a predetermined amplitude threshold (Fig. 4) and the movement of the robotic hand. We used above-threshold EMG amplitude of the external abdominal oblique and latissimus dorsi muscles to rotate the torso inward and outward during reaching and retrieving, respectively, to open and close the robotic hand. Since a duration of < 150 ms between intent and observation has been quantified as the response time to remain natural and enhance embodiment [12,30], we aimed to have a maximum response time < 150 ms.

**Fig. 4.**
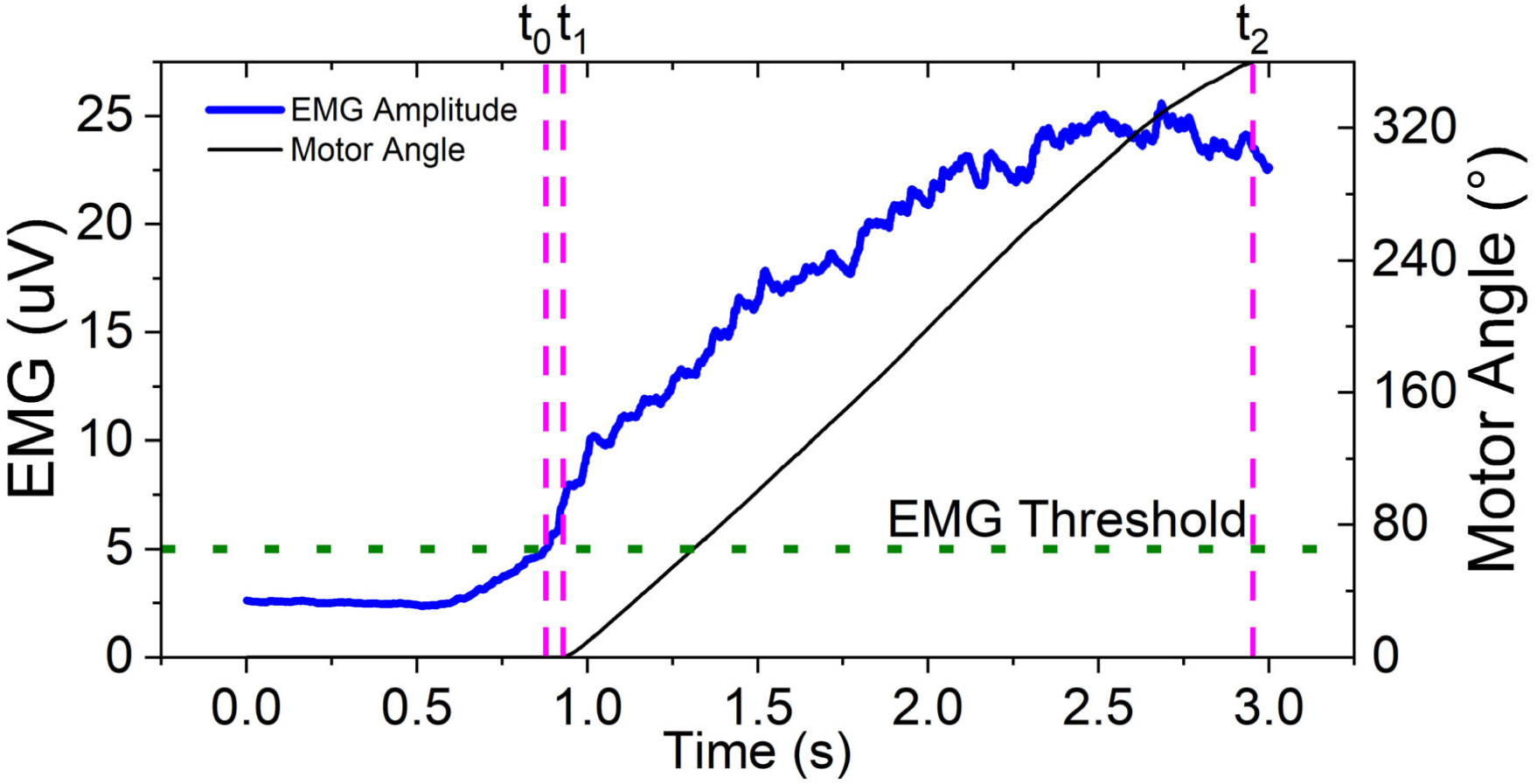
An example trial where a mimicked reaching (external abdominal oblique) task is performed and the robotic hand opens by sent motor commands. When the EMG amplitude of the torso muscle (blue line) exceeds the determined EMG threshold line (green dotted horizontal line), motor commands are sent to rotate the motors of the robotic hand. Critical time points for feasibility verification are labeled, t_0_ (the first detection of EMG amplitude exceeding the EMG threshold), t_1_ (the first observed increment to the motor encoder), and t_2_ (the final observed increment to the motor encoder).

### *B.* Detached Robotic Hand

We designed a simple and low-cost robotic hand (Fig. 3), placed distal to the hidden hand as if it is a functional extended hand (Fig. 2), with anthropomorphic, motorized actuation in digit flexion and extension, equipped with a speaker providing auditory feedback of digit motions. The robotic hand is a modified 3D print of the publicly available body-powered prosthesis CAD model (FlexyHand2, Thingiverse). The original design is modified by attaching a motor compartment to the wrist plate to house two DC Motors (78:1 Metal Gearmotor, 6V, Pololu #3465, Las Vegas, NV, USA), reduce stress on the connection between the wrist and palm by enlarging the width of the connecting bracket, and accentuate the extension of the little finger by rotating the metacarpophalangeal joint into abduction to be more visible to the participant in the pronated position. The robotic hand is 3D-printed using PLA material, and the joints are simulated by liquid silicone compounds (dragon skin 30, Smooth-On). It acts as an underactuated system in which a single motor simultaneously controls one direction, flexion or extension, of all 14-digit joints. The extension is actuated through a cable (Spectra parachute cord, Fire Mountain Gems, OR, USA) running from the fingertip through the palm and tied to the motor at the palmer side of the wrist. The flexion actuation is concentrated on the bottom of the palm where pairs of fingers (little and ring) and (index and middle) are connected through whippletrees to generate synchronized motion. A two-wire (Passive electronic buzzer, 2 terminal) speaker is incorporated near the robotic hand to enable auditory and visual modalities from a shared source location. It allows the participant to hear auditory feedback, associating a sound with a digit position.

The motion commands originate in custom-made software (LabVIEW, 2015, National Instruments, Austin, TX, USA) and are sent via serial commands to a microcontroller board (Arduino Mega 2560, Arduino, Ivrea, Italy) at 40 Hz, where velocity control is implemented through the motor shield (Dual TB9051FTG Motor Driver shield, Pololu, Las Vegas, NV, USA). The maximum motor speed is 100 rpm (Vmax), and a minimum speed of 25 rpm (25% Vmax) is required to move either motor to create motion in the fingers. The motion is controlled through a simple on-off control scheme, and displacement limits are set at the designated opened and closed orientations of the robotic hand. A motor encoder (Magnetic Encoder, 20 CPR, Pololu, Las Vegas, NV, USA) attached to each motor evaluates the motor position in real time. The motor position determines when motor commands should be halted and the pitch of the auditory feedback. The movement from the opened to closed position linearly adjusts the sound frequency from 300 Hz to 75 Hz, and vice versa.

The control of the system is shown with a simplified diagram for motion and pseudocode (Fig. 5). Motion is initialized when an orientation goal is set through the EMG state machine. If the EMG of the latissimus dorsi or external abdominal oblique is greater than their respective thresholds, a target orientation is delivered to the feedback loop. To minimize the error between the targeted orientation and the current orientation sensed by the motor encoder, motor commands are sent to both motors. Only one motor is activated by a command. For example, when minimizing the error to the flexion position, motor_1_ is set to on while motor_2_ is passive and driven by the rotation of motor_1_. This state is reversed when minimizing the error to the extension orientation. Motor commands are continually sent as long as an EMG signal exceeds its threshold and the error between the sensed orientation and the desired position is non-zero. If both EMG thresholds are simultaneously surpassed, no motion is produced. Well-defined thresholds eliminate the possibility of muscle co-contraction actuating the system and ensures the direction of motion matches the intention of the user.

**Fig. 5.**
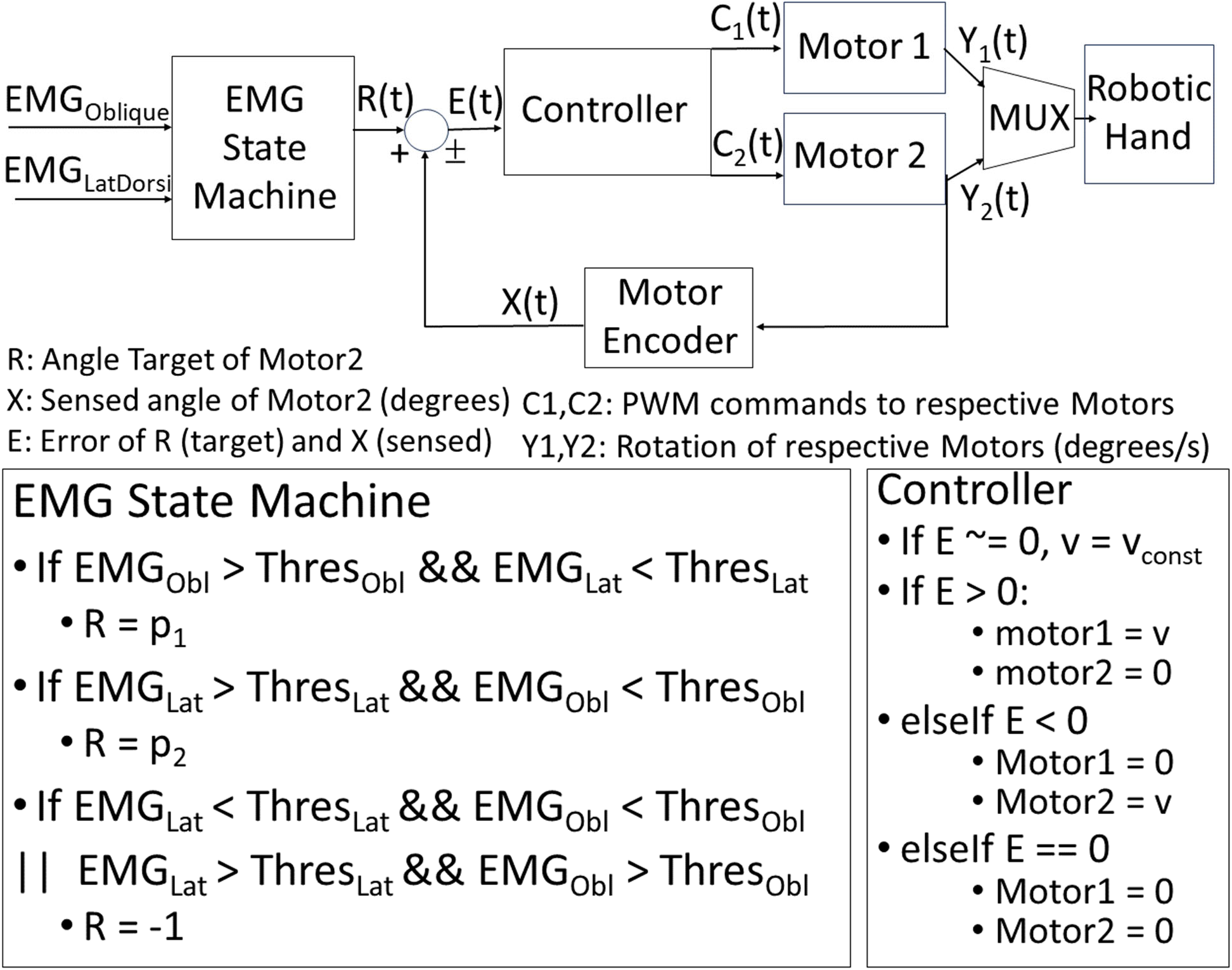
Control diagram of DC motors actuating robotic hand and associated pseudocode. The on/off control scheme of the robotic hand is enforced by a valid range of motor encoder values. Torso EMG above defined thresholds is necessary to generate motor commands.

### *C.* System to Translate Human Intent

Stroke survivors often have impaired limb mobility on the affected side, and they rotate the torso inwards during reaching and outwards while retrieving an object as compensation. The intent of reaching and retrieving is detected from EMG in the external abdominal oblique and latissimus dorsi, respectively. Surface EMG signals are amplified in a bipolar configuration (Z03-003, Motion Lab Systems, Baton Rouge, LA, USA), digitized at 1,000 samples/s (NI-PCI-6259, National Instruments, Austin, TX, USA), full-wave rectified, filtered (High-pass filter at 100 Hz, Butterworth order 3, exponential smoothing function tau = 0.3), and act as an on-off threshold for the robotic hand motion. Separate thresholds are determined in each user and set for the opening and closing of the robotic hand, sufficiently at a level where coactivation does not prompt the reverse motion. When the EMG amplitude is above the threshold, the microcontroller processes the non-zero velocity command and actuates the robotic hand motors (Fig. 4). A user would start opening the robotic hand as if reaching out to an object and holding the extended position for a predetermined duration (Fig. 2, *top*) before beginning to close the hand and retrieve the targeted object in the case of opening a door with a handle (Fig. 2, *bottom*).

### *D.* System to Translate Human Intent

Stroke survivors often have impaired limb mobility on the affected side, and they rotate the torso inwards during reaching and outwards while retrieving an object as compensation. The intent of reaching and retrieving is detected from EMG in the external abdominal oblique and latissimus dorsi, respectively. Surface EMG signals are amplified in a bipolar configuration (Z03-003, Motion Lab Systems, Baton Rouge, LA, USA), digitized at 1,000 samples/s (NI-PCI-6259, National Instruments, Austin, TX, USA), full-wave rectified, filtered (High-pass filter at 100 Hz, Butterworth order 3, exponential smoothing function tau = 0.3), and act as an on-off threshold for the robotic hand motion. Separate thresholds are determined in each user and set for the opening and closing of the robotic hand, sufficiently at a level where coactivation does not prompt the reverse motion. When the EMG amplitude is above the threshold, the microcontroller processes the non-zero velocity command and actuates the robotic hand motors (Fig. 4). A user would start opening the robotic hand as if reaching out to an object and holding the extended position for a predetermined duration (Fig. 2, *top*) before beginning to close the hand and retrieve the targeted object in the case of opening a door with a handle (Fig. 2, *bottom*).

In our human subject tests, participants were seated in a chair upright against a backrest (Fig. 2, *top right*). Since we were interested in the effect of cognitive engagement through neural activation, the torso was constrained by a harness to minimize torso motion, limiting the participant to static contraction. The right elbow and forearm of the participants rested on the table and were supported by a vacuum foam pad to stabilize the forearm. The biological arm of the participant was placed proximal to the robotic hand and hidden beneath a light cloth (Fig. 2, *bottom right*).

### *E.* Confirmation of System Functionality

We confirmed the functionality of the system by running it 50 times with and without human interaction. When an individual reaches and grasps a door handle, the metacarpophalangeal joint of the index finger travels ∼45° [22]. We aimed to replicate displacement in the robotic hand within 5% error, i.e., 45 ± 2.3°. To confirm it, the joint angle was recorded with an electronic single-axis goniometer (F35, Biometrics, Ladysmith, VA) placed over the metacarpophalangeal joint of the index finger of the robotic hand. The data were acquired at 1,000 samples/s (DataLink, Biometrics, Ladysmith, VA) and analyzed in custom software. The angle was zeroed when all digits were fully extended in the first trial. The final position was recorded at both ends after the angle did not deviate by 0.3° for 50 ms. The added goniometer mass reduced the distance traveled, given the same command. Considering the slower motions in stroke survivors than able-bodied adults and the facilitation of motion observation and motor imagery, we chose the target duration of opening and closing the robotic hand as 1.5-2 s. The desired duration required the motors to be set to an initial velocity of 35% Vmax to generate the 45-degree joint motions. The first sample of each trial was removed to eliminate delays from a prolonged idle state. The motion duration was defined from the encoder signals as the first observed change in position (t^1^) to the last observed change in position (t^2^) in Fig. 4. The system had to initiate movement in the robotic hand following EMG detection in an acceptable period to maximize opportunity for robotic embodiment. A threshold of 150 ms was set as the upper limit of the response time to be considered acceptable. The time between EMG detection of the torso muscles (t_0_) and the first observed change in position (t_1_) of the robotic hand quantified the observed response in Fig. 4.

### *F.* Feasibility of Human-Robot Interaction

To confirm the feasibility of using the system, an able-bodied adult (age range: 26-30 years old) was recruited. The participant controlled the detached robotic hand through contracting the external abdominal oblique and latissimus dorsi muscles. They contracted each muscle independently to determine thresholds that were at a comfortable level and greater than the amplitude during unintended coactivation. The task included 1) increasing and maintaining the contraction level of their external abdominal oblique until the robotic digits extend and reach full extension, 2) holding the contraction level for ∼1.5 s, and then 3) increasing and maintaining the contraction level of their latissimus dorsi until the robotic digits flex and reach full flexion. The participant was asked to observe the robotic hand motion, listen to the auditory feedback, and add kinesthetic imagery. Kinesthetic imagery is imagining the feeling of their muscles and the external environment as if they were performing the task with their biological hand. After 10 trials, the participant rated their sense of embodiment of the robotic hand by marking a point on a 10-cm visual analog scale (10 cm horizontal line without ticks on a paper VAS), with 0 being no embodiment and 10 being strong embodiment. The sense of embodiment was described to the participants as how well 1) they felt connected to the robotic hand through observation and imagery (if required in the task) and 2) the motion of the robotic hand mimicked their intent during its control with their torso muscle activation.

### *G.* Feasibility to Test Biological Hand Function

The system was further developed to enable the testing and training of the biological hand while the torso muscles interact with the detached robotic hand. It will help future research in assessing the effect of using the developed system. The system is equipped with a hi-pitch sound (440 Hz) that provides a cue to initiate a certain task when the robotic digits reach a specified position. A set of random encoder values within the range is implemented. The system is equipped with a sensor (model 31/34, Honeywell, Charlotte, NC) below the biological index finger to assess its flexion force). The sensor output is sampled at 1,000 samples/s (NI PCI-6259, National Instruments, Austin, TX, USA). The participant performed biological hand tests concurrently with three tasks (robot interaction, motor imagery, and resting) to examine the feasibility of adding biological hand tests to robot interaction. The three tasks are explained after the explanation of the biological hand tests below.

To examine the feasibility of assessing the biological finger performance, a reaction time test and a maximal voluntary contraction (MVC) test were performed with the right index finger during the three tasks in the able-bodied participant. During each task, the participant exerted finger flexion forces with their index finger as soon as they heard an auditory cue for the reaction test or as much as possible for the MVC test. In the reaction time test, there was no instruction on the amount of force or rate of force development. In the pronated position, the participant rested their right index finger on a force sensor. They kept their arm and hand muscles relaxed until they responded to the cue by pressing on the force sensor with the index finger. The participant performed 10 trials for each test during each task. The biological hand tests were performed concurrently with robot interaction (see section *E*) and motor imagery. The resting condition was also included as a reference of performing the tests without involving intentional effort to perform another task concurrently. The hand test with robot interaction or motor imagery task is a dual task requiring divided attention, whereas that in the resting condition is a single task in which participants were able to focus their attention on the finger motor execution. Motor imagery was performed because it is the standard mental practice in stroke rehabilitation. The participants mentally visualized a task in which they would reach out to a door handle, grasp it, and pull it to open the door. After completing the reaching portion, the participant began the pulling portion, keeping their eyes open and responding to the audio cue with their index finger for the biological hand tests. In the reaction time test, the reaction time was defined as the duration between the cue and the force onset, the maximum slope of the fitted line before peak force was reported as the maximal rate of force development, the peak reaction force was determined as the maximum peak force, and peak reaction FDI EMG amplitude was reported as the maximum FDI EMG in the reaction time test. The obtained values were averaged across the first 9 acceptable responses. In the MVC test, the maximal peak force across three trials was determined as MVC force. The determination of these variables was performed within a custom script (MATLAB).

### *H.* Applicability to Stroke Survivors

After the feasibility of the tasks and functionality of the system were confirmed with an able-bodied individual, we assessed the applicability to stroke survivors and observed the acute effects of this robot interaction on their affected hand function. We recruited five adults with mild or moderate hemiparetic upper-limb impairment because of a stroke (23-69 years old (mean: 53.2 years old, 2F, 3M). All participants had volitional control over the flexion of their index finger on their affected hand. Some of the participants experienced spasticity in their hand and fingers that required manual manipulation to extension with their other hand to maintain comfort while resting on the force sensor. Those who did not have spasticity had some level of muscle weakness varying from the distal and proximal muscles. All the participants completed the same three tasks described above (resting, motor imagery and interaction with robot) with and without biological hand function. The sense of embodiment was recorded after the participant completed the robot interaction task without biological hand function. The related biological hand function variables were compared as raw values and then normalized to the resting condition to assess the relative changes in participants with various impairments. A paired t-test was performed for each variable across the post-stroke participants. A Shapiro-Wilk test was performed on each data set to ensure normality was satisfied considering the low sample size from this exploration. For comparisons that were statistically significant the normality, effect size (Cohen’s D) and p values are reported. The alpha value of 0.05 was used for statistical significance.

## Results

### System functionality and feasibility

The range of motion capabilities of the robotic hand was verified through 50 trials (Fig. 6). On average, the robotic hand was oriented at 0.84° (extended) and 42.10° (flexed) at the end of its motion. Across the 50 trials (Figs. 6a & 6b), the duration from the closed to the opened position, during which the digit inertia load is overcome, was 1.85 ± 0.045 (mean ± SD) s. The duration from the opened to closed position was slightly shorter, 1.54 ± 0.032 s. The system responded consistently and within our desired bounds for both directions of movement, with response delays of 97.2 ± 5.76 ms for opening and 47.8 ± 3.43 ms for closing.

**Fig. 6.**
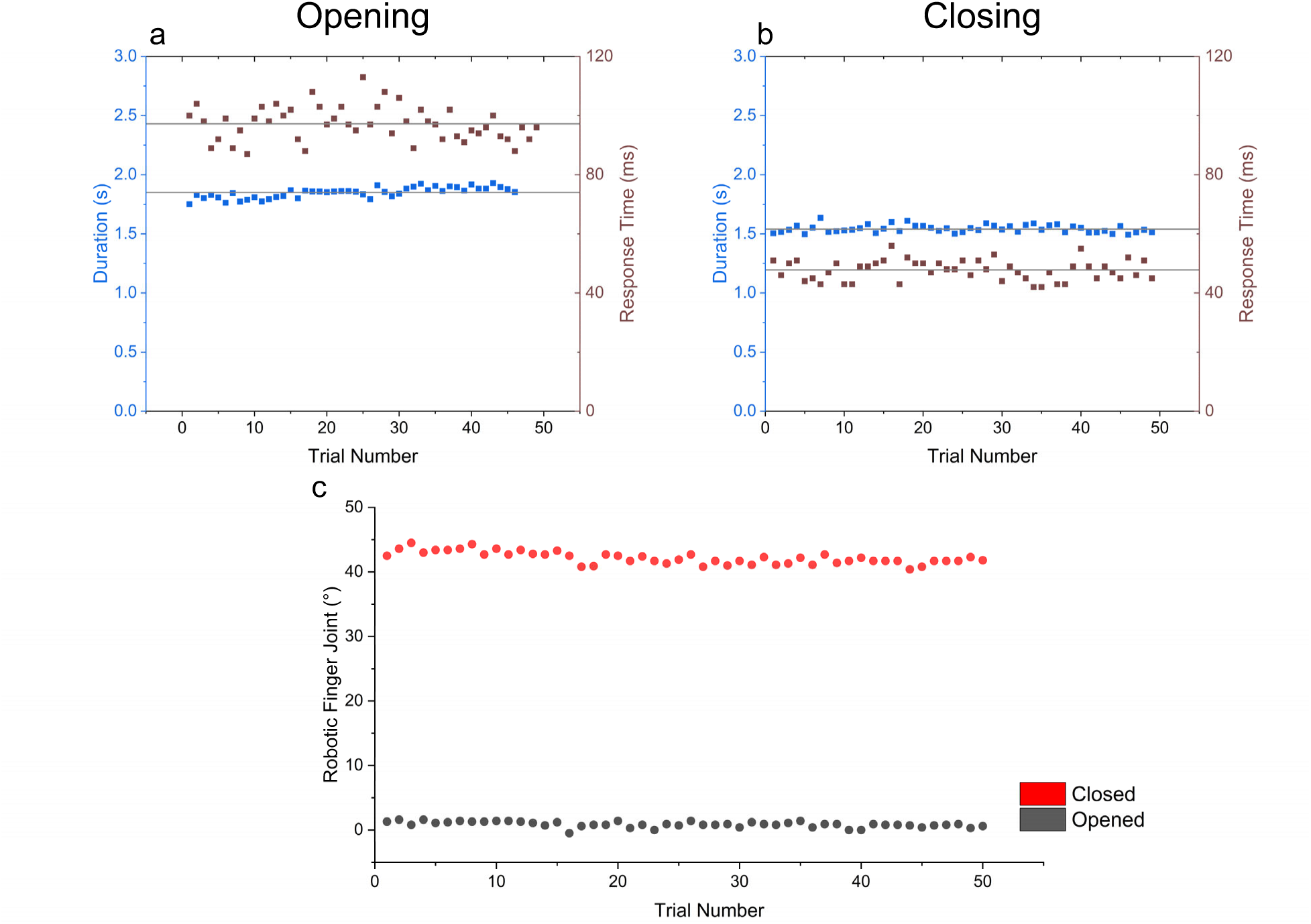
Mechanical characteristics of the robotic hand, including compiled durations for opening (a, blue) and closing (b, blue) in the chosen velocity configuration. The response time for opening (a, brown) and closing (b, brown) the hand in response to torso EMG. The final orientation (c) of the metacarpophalangeal joint of the robotic index finger.

We first recruited an able-bodied adult to confirm the feasibility of the system and test the structure of the tasks we intended to perform with post-stroke participants. The able-bodied participant did not have any issue in controlling the robot with torso muscle contractions after a short practice. The embodiment score after 10 trials of robotic interaction was 5.4 out of 10. Subsequently, biological hand tests during robot interaction and other tasks were performed, including the reaction time test and the MVC test. In all trials, the participant was able to respond to each random auditory cue by discerning it from the continuous auditory feedback on the robotic digit movement. After sufficient practice, we confirmed that the participant was able to complete each task smoothly and with a full understanding of the instruction and expectations of each task. We confirmed the appropriate recording of force profile (Fig. 7) for subsequent data analysis across conditions.

**Fig. 7.**
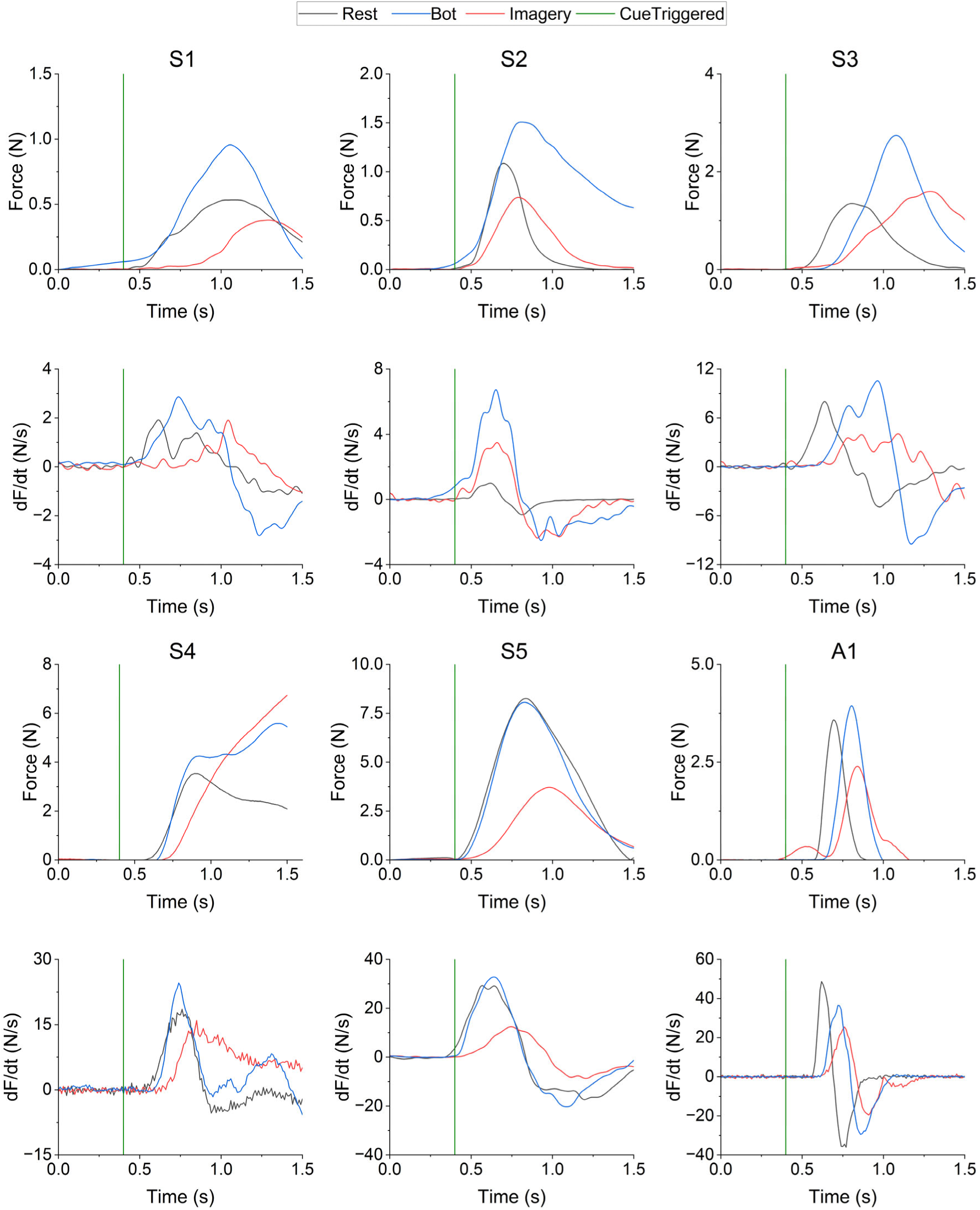
The force response (first and third rows) and rate of force development (df/dt, second and fourth rows) of the index finger during the reaction time test are trigger-averaged to the onset of the audio cue (green vertical line) in six participants (S1-S5: Stroke; A1: Able-bodied). The data during the resting (black), robot interaction (Bot, blue) and motor imagery (Imagery, red) tasks are presented.

**Fig. 8.**
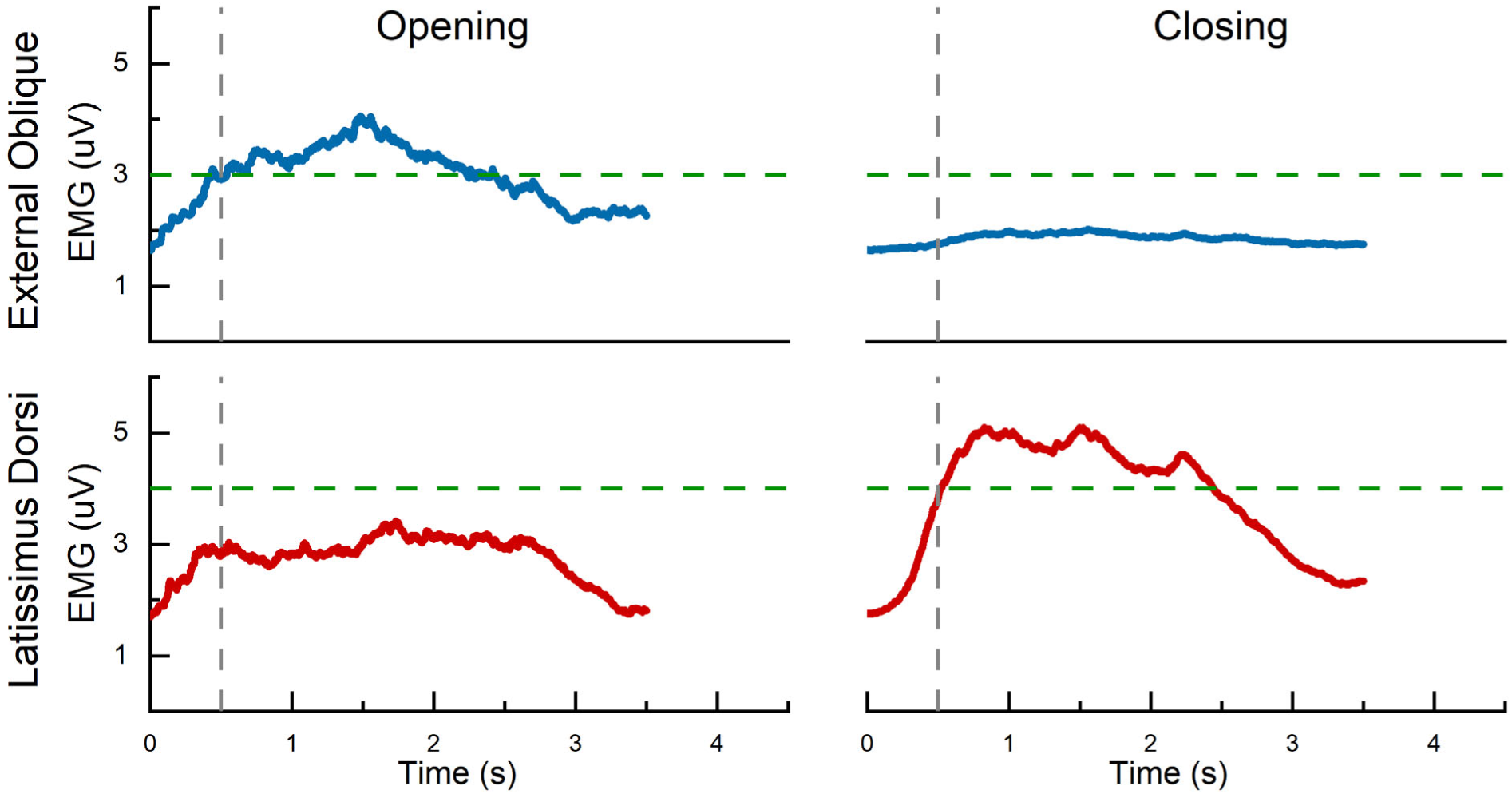
The EMG amplitude of the driving torso muscles of a stroke survivor for opening (left column) and closing (right column) the robotic hand. The horizontal lines (in green) show the threshold to trigger the corresponding robotic hand motion and the vertical line (in gray) the time when the active muscle crosses the defined threshold. The external abdominal oblique (upper row, in blue) was used for opening, and the latissimus dorsi (lower row, in red) was used for closing the robotic hand. The appropriate thresholds were determined so only one muscle passed the threshold for each robotic motion despite coactivation.

### Applicability to stroke survivors

The external abdominal oblique and the latissimus dorsi may exhibit coactivation during the torso muscle activation to mimic reaching and grasping during the robot-human interaction, especially in stroke survivors. Nonetheless, we were able to determine a unique threshold for the corresponding muscle for both reaching and grasping in each post-stroke participant so that the coactivation of the secondary muscle remains below the threshold. An example is shown for one opening and closing trial (Fig. 7). All participants were able to surpass and maintain the EMG thresholds to control the robotic hand smoothly and repeatedly. The embodiment score after 10 trials of robotic interaction was 6.5 ± 2.0 (3.3-8.8) across post-stroke participants.

Biological hand tests during robot interaction and other tasks included the MVC test and the reaction time test. In general, the MVC force tended to be lower and the reaction time tended to be longer during both dual-task conditions (i.e., robot interaction task and motor imagery task) compared with the resting condition (single task) (Fig. 9). Nonetheless, the MVC force during robot interaction was greater compared with those during motor imagery in four of five participants and comparable in the fifth (Fig. 9). The data did not satisfy the normality test (normality: p=.006, Cohen’s D: .657, p = .108) or significance. We did not observe any trend in the reaction time between robot interaction and motor imagery conditions. While the normality test was satisfied for reaction time, there was no significant difference (normality: p=.112, Cohen’s D: .173, p = .36) between the robot interaction and imagery tasks.

**Fig. 9.**
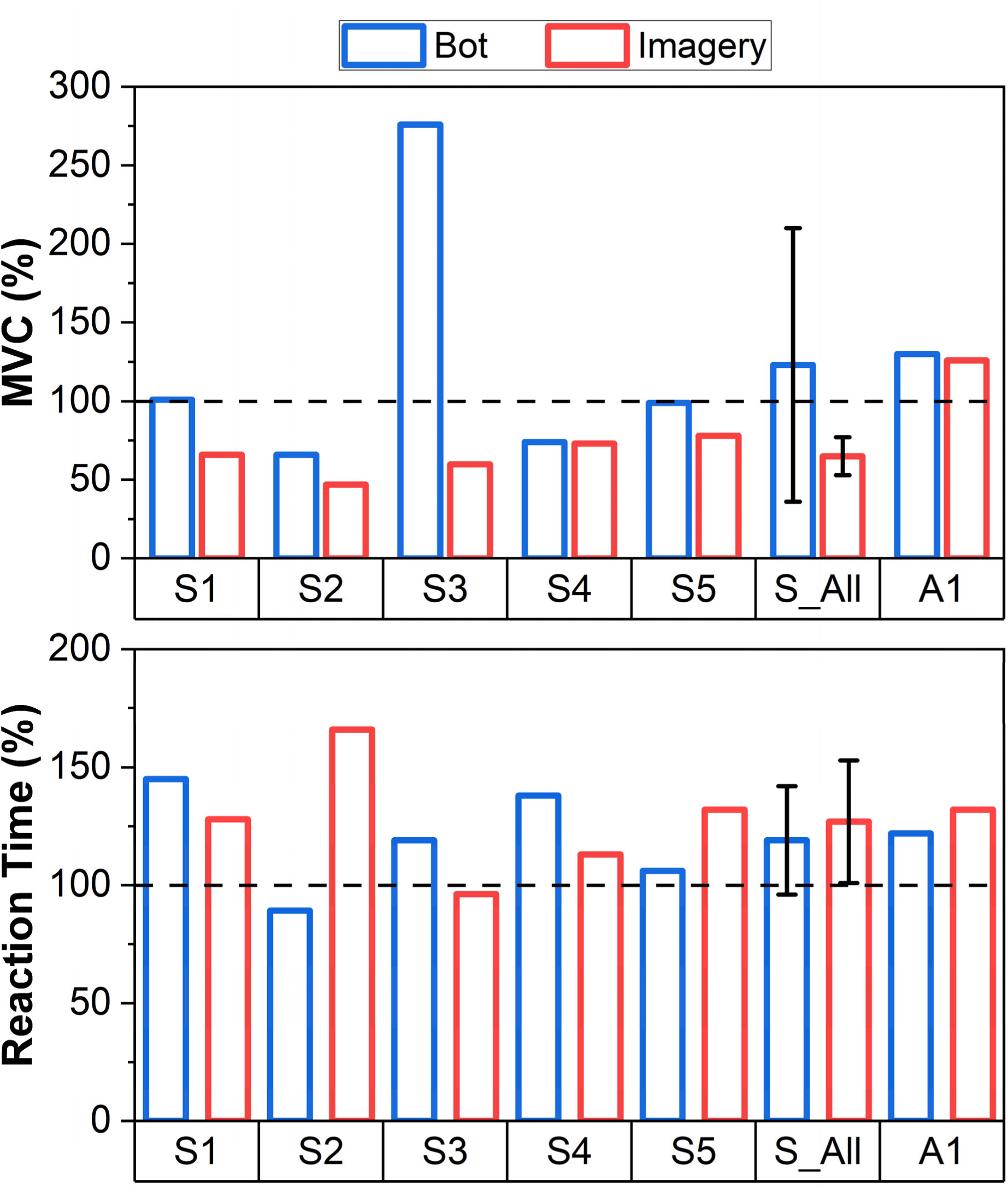
Maximum force during maximal voluntary contraction (MVC) test and reaction time during the reaction time test in stroke (S1-S5) and able-bodied (A1) participants. The values presented are normalized to the respective rest value for each participant. The robot interaction (Bot, blue) and motor imagery (Imagery, red) tasks are presented for each participant, and the average across the five stroke participants (S_All) is also presented. A dashed horizontal line is added to easily visualize the comparison to the rest condition. Data for an able-bodied participant are also presented. *, P < 0.05

While the participants were not instructed on the amount of force or the rate of force development during the reaction time test, the profiles were variable depending on the task. The trigger averaged force response and the derivative of force for each condition are shown in Fig. 7. Compared with the resting, the resultant peak reaction force (Fig. 10, top) and the maximal rate of force development (Fig. 10, middle) were higher during robot interaction in most participants. In four out of five participants, the peak reaction force was higher compared with the resting (by 36% to 136%). The fifth participant only showed a comparable value. On average, the peak reaction force was higher by 65% across post-stroke participants. Additionally, the peak reaction force was significantly greater (normality: p=.996, Cohen’s D: 1.09, p = .036) while interacting with the bot compared to the imagery task. The maximal rate of force development was higher in all post-stroke participants during robot interaction compared with rest. On average, it was higher by 26% (7 to 48%). This consistent increase during robot interaction was in contrast with the consistent decreases during motor imagery in all post-stroke participants (−4 to −49%, −22% on average). The rate of force development was also significantly greater (normality: p=.420, Cohen’s D: 6, p < .001) while interacting with the robot compared to the imagery task. Compared to the resting condition, the peak EMG amplitude increased during the robot interaction in four out of five post stroke participants (Fig. 10, bottom). The other participant showed a comparable value. On average, the peak EMG amplitude was higher by 66% across post-stroke participants. The peak EMG amplitude during the task was also significantly greater (normality: p=.965, Cohen’s D: 1.24, p = .025) during the robot interaction than during the imagery task.

**Fig. 10.**
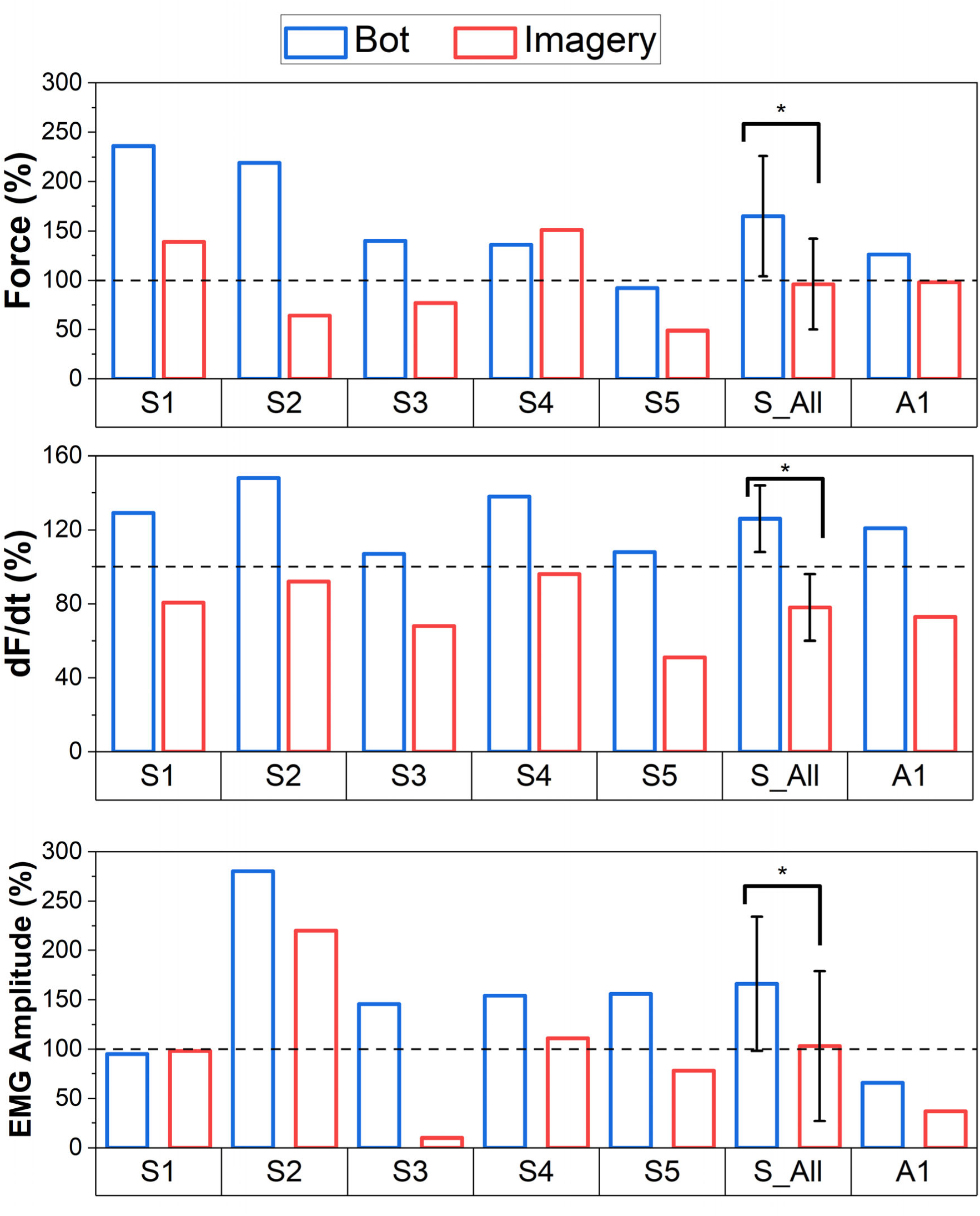
The maximum force response, maximum rate of force development (dF/dt) and maximum EMG amplitude in the first dorsal interosseus muscle during the reaction time test in stroke (S1-S5) and able-bodied (A1) participants. The values presented are normalized to the respective rest value for each participant. The robot interaction (Bot, blue) and motor imagery (Imagery, red) tasks are presented for each participant, and the average across the five stroke participants (S_All) is also presented. A dashed horizontal line is added to easily visualize the comparison to the rest condition. Data for an able-bodied participant are also presented. *, P < 0.05

## Discussion

We envisioned a novel neurorehabilitation system that utilizes the synergistic proximal muscle activations associated with reaching, opening, and closing a hand to grasp and retrieve an object. It involves active control and observation of the opening and grasping actions of a detached robotic hand. Accordingly, we developed a simple system to produce the motion in an anthropomorphic robotic hand that performed the tasks within the targeted specifications at a minimal cost.

The intended duration of the robotic hand for both extension and flexion movements in feasibility testing was 1.5-2 s, which is the middle ground for able-bodied and moderately impaired individuals. Since the physical system did not compensate for gravity, the closing motion was assisted by gravity and completed its trajectory in a shorter duration (1.54 s) than the opening motion (1.85 s), which is opposed by gravity. The effects of gravity are evident because the same velocity command is sent to initiate the motion in both directions. Although we acknowledge the potential improvement in precision and repeatability by compensating for gravity, the results show that the system performed within the 1.5-2 s duration we defined, and no perceivable differences were noted during the operation of the system.

The intended range of motion of the robotic hand within the constraints of grasping a door handle was ∼45°. The mass and size of the goniometer altered the motion of the robotic hand depending on the location of attachment. Specifically, the proximal joint was selected to reduce the observed moment when the goniometer is attached to the more distal regions. When attached distally, the goniometer significantly altered the observed motion, as a less smooth trajectory was observed. The purlicue (the skin region between the thumb and forefingers) was therefore chosen as the attachment region as there is no interference from the actuating tendons and its proximity to the base of the hand. Despite our considerations, the goniometer still affected the motion slightly, the index finger did not return to the zero position during opening, remaining ∼1° flexed. The average resting orientation of the metacarpophalangeal joint of the robotic index finger during flexion was measured to be 42.1°, and the average extended orientation was 0.84°. The observed orientation of the robotic hand reasonably replicated that of the biological hand, while the cylindrical shape of the door handle enabled a simple motion as all four fingers achieved their full grip synchronously. The shape of the door handle limited the range of motion that we intended to produce, but the measured closed orientation (42.1°) visually appeared as a fully closed hand to the participant. With the challenges of measuring each joint of the robotic hand, the measured data from the metacarpophalangeal joint of the index finger supports the repeatable and representative opening and closing motions of the hand during reach and grasp.

The desired response time between user intention (i.e., EMG onset) and robotic hand motion was < 150 ms as determined from a control perspective [12]. It is also known that the degree of embodiment is significantly decreased if a time delay exceeds 200 ms [30]. Since the initiation of the robotic motion needed to be reasonably aligned with the recognition of user intention to increase the opportunity for intuitive control and effective embodiment, the more constraining 150 ms was selected as our target. The experimental configuration with a 10-ms command interval limited the variability between trials substantially, and no trial had a response time greater than 120 ms. The observed response time was shorter than the reported time in other studies with exoskeletons (e.g., 130 ms [31] and 153 ms [33]). The longer response time in the opening was because the system remains simple and does not compensate for gravity, requiring a greater torque to initiate movement in opening the closed robotic hand. The response time of our system can be further reduced by compensating for gravity.

The primary purpose of developing the robotic hand was to create a simple and low-cost system that can be used to demonstrate and test the proof of concept of a new rehabilitation paradigm with a detached robot and possibly implemented without a high technical or financial burden at various sites. Without trying to replicate the complexities of the full functions of a human hand, the choice to replicate the opening and closing motions allowed for simplification. The metacarpophalangeal joints of the five digits are tied together, and through 2 DC motors a unified opening and closing motion representing 1 DOF is achieved. Although a single object (door handle) was used throughout testing in the current study, the motion of the human hand is object dependent. A form of adaptive grip was created through the pathing of the tendons, and the four fingers would move synchronously until an object or length of the cabling limits the motion. For grasping a ball, for example, the longer fingers may continue to grasp around the ball while the little finger is restricted. Intricate movements of the hand were not replicated, including translation of the hand. While each added feature may further improve human embodiment, the simple design can validate the function of the system without requiring complicated control or overwhelming experimenters.

For the low cost, we aimed to keep the total cost <US$150. The robotic hand was 3D-printed from PLA material, and the total volume of functional and support material was ∼400 g. A roll of 1 kg PLA can be purchased for <US$25. The tendons of the robotic hand were constructed through Spectra parachute cord, totaling approximately US$10 for a roll. The actuation and control components totaled approximately US$110 for two motors, a motor shield, two encoders, and a microcontroller. While this system will not be immediately deployed to rehabilitation clinics, the minimal cost would allow easy access.

Our unique paradigm was to use the synergistic torso muscle activations, which often accompany reaching and retrieving movements, especially in stroke survivors, to control a detached robotic hand. To focus on the contribution of neural activity and minimize the involvement of human movements, the upper body was constrained to the chair. In this posture, the feasibility of robot-human interaction was tested with and without a biological hand test. With a few practice and adjustment trials, we were able to determine independent thresholds in the EMG amplitude for the external abdominal oblique and latissimus dorsi for controlling the opening and closing motions of the detached robotic hand, respectively, in both able-bodied and post-stroke adults. The embodiment scores of 5.4 in able-bodied and an average score of 6.5 in post-stroke participants out of 10 indicate they had a good sense of having the detached robotic hand as part of their body through only 10 trials of controlling, observing, and listening to its movement and associated sounds.

Previous studies questioned the effectiveness of a 2D virtual hand in engaging the mirror neuron system during action observation and motor imagery to the same extent seen when interacting with a 3D model (physical or virtual) [13,17]. Although a 3D virtual reality system has shown to have similar engagement to a 3D object in the physical world, the constraints of the experimental design may limit the practicality of its implementation. For example, suppose transcranial magnetic stimulation (TMS) is incorporated for concurrent measurements or therapy. In that case, the magnetic effects and required location (the top of the skull) of the TMS coil can affect the usability of a headset for a 3D virtual reality. Therefore, this 3D robotic hand system was developed to take advantage of increased embodiment without disrupting associated experimental approaches. Since the process of converting the 3D printing files to a 2D virtual hand is straightforward and recent physics simulators are advanced and realistic, adoption and examination of a 2D virtual hand compared with the 3D physical system in future studies are feasible. It is important to evaluate if the improved technology bridges the previously observed differences in engagement and to enable an alternate, yet suitable approach.

In addition to evaluating the functionality of this system, we investigated the feasibility of concurrently testing the motor function of the biological hand for future application with this developed system. In the able-bodied and post-stroke participants, the randomly provided auditory cue for the test was discernable from the ongoing sounds corresponding to the robotic hand movements. The execution and observation of the concurrent hand motor tests strengthen the feasibility of using the developed system in various ways. Since dual-task executions require more attentional resources compared with single-task executions, we had expected a degradation of intended performance in the biological hand test with motor imagery or robot interaction compared with the single-task execution at rest, which were observed as longer reaction time and lower MVC force. In the reaction time test, in contrast, the greater peak reaction force, maximal rate of force development, and FDI EMG amplitude during robot interaction compared with the resting in most post-stroke participants are exciting observations that demonstrate the applicability of facilitating the neural activation for hand function in stroke survivors. It is noted the one participant (S5) who reported the lowest embodiment score (3.3) was the same individual who did not generate a larger reaction force during robot interaction compared to rest. The participant’s limited perception of embodying the robotic hand may be involved in the limited enhancing effects. Still, these variables during robot interaction were greater compared with motor imagery in this participant. Additionally, significantly greater peak reaction force, maximal rate of force development, and FDI EMG amplitude during robot interaction compared with motor imagery, i.e., within dual tasks, across all post-stroke participants suggest the facilitating potential of controlling the detached robotic hand with synergistic torso muscles.

In the present study, we aimed to create a novel system that would support the unique inquiry toward the potential effects of combining synergistic torso muscle activation with active control of a robotic hand to assist stroke rehabilitation. The created system proved capable of producing motion akin to biological humans in terms of the duration and range of motion. Like humans, the motion was consistent from trial to trial, as observed for the duration and range of motion, and response time to torso muscle effort. The reduction of the system to one degree of freedom allowed for the above metrics to be met and yet, an affordable final cost. In addition to the duration we selected for feasibility testing, the system supported a range of durations to present future opportunities to explore alternative target populations. The feasibility of the system and applicability to clinical populations were demonstrated with able-bodied and post-stroke adults with impaired hand function. Notably, increased reactive force and hand muscle activity were observed in most participants. The study thus produced an operational system that is ready to be used in future studies to systematically investigate the effect of synergic torso-muscle control of a detached robotic hand in various populations.

## Data Availability

All data produced in the present study are available upon reasonable request to the authors

## Acknowledgement

The authors thank Jun Ueda for providing constructive feedback on an earlier version of the manuscript.

## Notes

**Funding:** This work was supported by the National Institutes of Health/National Institute of Neurological Disorders and Stroke (1R21NS118435-01A1).

### Competing Interest Statement

The authors have declared no competing interest.

### Clinical Trial

NCT04962698

### Funding Statement

This study was funded by the National Institutes of Health/National Institute of Neurological Disorders and Stroke (1R21NS118435-01A1).

### Author Declarations

IRB of Georgia Institute of Technology gave ethical approval for this work (H21154).

### Summary of Updates

The author name of Minoru Shinohara has been corrected by removing the middle initial N. The name of affiliation of Andrew J Butler has been corrected to School of Health Professions.

